# Association of Life’s Essential 8 with Cardiovascular Events and Mortality: The Cardiovascular Disease Lifetime Risk Pooling Project (LRPP)

**DOI:** 10.1101/2023.08.08.23293855

**Authors:** Hongyan Ning, Amanda M. Perak, Juned Siddique, John T Wilkins, Donald M. Lloyd-Jones, Norrina B. Allen

## Abstract

**Background:** The American Heart Association (AHA) recently launched updated cardiovascular health (CVH) metrics, termed Life’s Essential 8 (LE8). Compared to Life’s Simple 7 (LS7), the new approach added sleep health as an eighth metric and updated the remaining 7 metrics. Whether the updated LE8 score outperforms the original LS7 score in predicting cardiovascular disease (CVD) is not known. We examined the association of LE8 scores with CVD, subtype CVD events and all-cause mortality.

**Methods:** We pooled individual-level data from 6 contemporary US-based cohorts from the Cardiovascular Lifetime Risk Pooling Project (LRPP). Total LE8 score (0-100 points), LE8 score without sleep (0-100 points), as well as prior LS7 scores (0-14 points), were calculated separately. We used multivariable-adjusted Cox models to evaluate the association of LE8 with CVD, CVD subtypes, and all-cause mortality among younger, middle, and older aged adult participants. Reclassification was defined based on the concordant/discordant categories of LS7 and LE8 scores quartile rankings across age groups.

**Results:** Our sample consisted of 32,896 US adults (7836 [23.8%] Black; 14941 [45.4%] men) followed for 642,000 person-years; of whom 9,391 developed CVD events. Each 10-point higher overall LE8 score was associated with 23-40% lower CVD risk across age groups. Reclassification of CVH from LS7 to LE8 was related to heath behaviors as well as health factors and was significantly associated with CVD risk.

**Conclusions:** These findings support the improved utility of the LE8 algorithm for assessing overall cardiovascular health and future CVD risk.

## Introduction

Cardiovascular disease (CVD) remains the leading cause of death globally, accounting for 32% of all deaths^1 2^. Smoking, obesity, poor diet, sedentary lifestyle, insufficient sleep, hypertension, dyslipidemia, and diabetes have been identified as major CVD risk factors ^3–5^. The American Heart Association (AHA) has increasingly focused on primordial prevention to promote cardiovascular health (CVH) and reduce the burden of CVD and its risk factors ^6^. In 2022, AHA defined updated metrics, termed Life’s Essential 8 (LE8), to measure and monitor CVH and promote improvements in individual and population health^7^. The new metrics represent a modification of the earlier Life’s Simple 7 (LS7) metrics for CVH and includes sleep health as an eighth CVH metric. ^4^ LE8 includes 4 health factors (body mass index, non-high density lipoprotein cholesterol, blood glucose, and blood pressure) and 4 health behaviors (diet, physical activity, nicotine exposure, and sleep) ^7–9^. Recent studies showed that overall LE8 CVH scores are suboptimal in US adults, with overall mean score of 64.7 out of 100, and less than 20% of US adults achieve high CVH status (LE8 score ≥80)^9 10^.

The construct of CVH, and the ability to quantify it, have proven to be useful tools to promote overall health and healthy longevity. Numerous studies have shown that higher LS7 scores are associated with a multitude of favorable health outcomes, including markedly lower risks for cardiovascular events and death across the life course ^5, 11–13^. To date, no studies have evaluated the associations of the new updated LE8 score with cardiovascular outcomes and death through the life course, likewise LE8 sub-scores of health factors and behaviors have not been examined separately. Furthermore, although LE8 and LS7 scores are highly correlated, differences were observed between these two scores attributable to the addition of the new sleep metric and the distinct scoring systems ^9, 12^. The new LE8 scoring system for each metric ranges from 0 to 100 points (with overall LE8 score of 0 to 100 representing the mean of all metric scores) whereas the LS7 score ranges from 0 to 2 for each metric (with an overall range of 0 to 14 points representing the sum of all metric scores). Whether the broader range of the LE8 algorithm captures more inter-individual differences remains an open question. Finally, the significance of downward or upward reclassification of CVH status between the LS7 and LE8 scoring algorithms has not been examined.

Using data from the Cardiovascular Lifetime Risk Pooling Project (LRPP), we investigated the association of the LE8 score and its health factor and behavior sub-scores with cardiovascular events across young, middle, and older US adults. We hypothesized that LE8 scores would be strongly associated with cardiovascular events and death and would provide more granular description of CVH status and better discrimination of related CVD risk than the LS7 score. In addition, we hypothesized that reclassifying an individual’s risk from LS7 to the more nuanced LE8 definition would have statistical significance in differentiating and correctly classifying the participants with their predicted CVD risk.

## Methods

### Study sample

The Cardiovascular Lifetime Risk Pooling Project (LRPP) is an aggregation of 22 community- and population-based U.S. cohort studies^14^. For this study, we included participants from 6 contemporary cohorts: the Atherosclerosis Risk in Communities Study^15^, Cardiovascular Health Study^16^, Coronary Artery Risk Development in Young Adults Study^17^, Framingham Heart Study^18^, Framingham Offspring cohort^19^, and Multi-Ethnic Study of Atherosclerosis^20^. Dietary and physical activity data have been harmonized and published previously ^21^, therefore the baseline examination data in each cohort was defined as the first exam in which both diet and physical activity along with the most risk factor metrics were concurrently measured. Data were collected between 1985 and 2002, and the most recent follow up occurred up through August 30, 2020. This study was approved by the Institutional Review Board at Northwestern University. Written informed consent was obtained from all participants for initial data collection in each original cohort. For this analysis of deidentified data, specific consent was not required.

### CVH metrics

The LE8 score is composed of 4 health factors (body mass index (BMI), blood glucose, lipid, and blood pressure) and 4 health behaviors (smoking, physical activity, diet, and sleep). The definitions and levels of each component are described in **Supplemental Table 1,** consistent with AHA recommendations ^7, 9^. Each of the metrics is scored on a scale of 0 to 100, and the overall LE8 score is the average value of all the component scores (range, 0-100). Based on the two domains of LE8 metrics, LE8 health factor and LE8 health behavior scores were created. For comparison with LS7 scores, we also calculated LE8 score without the sleep metric (i.e. the average score 0-100 from the other 7 metrics). LS7 is composed of 7 metrics (no sleep), and each metric is scored as 0 to 2, and the overall LS7 score is the sum of the scores of all the 7 metrics (range, 0-14) as described in **Supplemental Table 2** ^8, 22^. The definitions and scoring system differ in LE8 and LS7. For example, in the LE8 algorithm non-HDL cholesterol level is used instead of total cholesterol, and scores are reduced for exposure to second-hand smoke and if risk factor levels were measured on treatment, such as antihypertensive, hypoglycemic, or lipid-lowering medications.

Detailed data ascertainment methods are available in the original design publications from each study. BMI was calculated as the weight in kilograms divided by the height in meters squared. Lipid, BP, fasting glucose and HBA1c were measured according to standard protocols. Nicotine exposure was determined based on self-reported tobacco use and questionnaire responses. Diet was assessed by food frequency questionnaires and linked to the Dietary Approaches to Stop Hypertension (DASH) score ^23^. Physical activity was assessed by different physical activity questionnaires used within each cohort and minutes per week of moderate- or vigorous-intensity physical activity were calculated. Sleep health was assessed using self-reported information about sleep duration. Additionally, information regarding education level, prior CVD history, and use of blood pressure-lowering, glucose-lowering, and lipid-lowering medications was obtained. Missing CVH metrics and covariates were imputed by multivariate imputation by chained equations (MICE) (for full details, see supplementary methods).

### Outcome Ascertainment

The primary outcome was major CVD events, defined as a composite of the first event of incident myocardial infarction, stroke, heart failure, or CVD death. Major CVD events were adjudicated using similar methods in each cohort included in the analysis ^14^. Methods for mortality ascertainment included contact with family and employers, and matching to Social Security Administration files, the National Death Index, and National Death Index-Plus service. Mortality ascertainment for all cohorts has been essentially complete. We excluded participants with a prior CVD event, and follow-up time was censored at the time of a first CVD event, death from other cause, or end of follow up data (ie, last known date alive).

### Statistical Analysis

Participants were divided into 3 index age groups based on age at baseline assessment: young (20-39 years), middle-aged (40-59 years), and older (60-79 years). All analyses were conducted separately by index age groups. Baseline characteristics of the participants are summarized as the mean (SD) for continuous variables and percentages for categorical variables. Multivariable Cox regression models were used to examine the association of the baseline LE8 score with CVD, subtypes of CVD and death. All models were adjusted for age, gender, race/ethnicity, and education level. For each CVH metric, we used Poisson regression models to calculate crude incidence rates (95%CI) of CVD across all the strata of LS7 and LE8 scoring, separately. To quantify and compare the distribution of LS7 and LE8 scores, we categorized the participants into four groups according to the age-specific quartiles (<25^th^ percentile, 25th percentile to median, median to 75th percentile, and >75th percentile) of LS7 and LE8 score separately. The agreement between the age-specific quartile ranking of these two scores was assessed using Cohen’s Kappa statistic. Concordant/discordant categories were derived based on the matched quartiles of LS7 and LE8 scores across index age group. The individuals were classified as concordant if LS7 and LE8 score had the same quartile ranking; otherwise, they were classified as discordant (i.e., reclassified). Upward/downward reclassification was defined with higher/lower LE8 quartile ranking, respectively, relative to the LS7 quartile within the index age group. A p-value <0.05 was considered significant, and SAS version 9.4 (Cary, NC) was used to conduct all analyses.

## Results

A total of 32,896 participants were included from the 6 cohorts in this analysis (mean [SD] age: 53.9 [4.2] years, 54.5% women; 23.8% Black; 55.3% with some college and higher education). They contributed 642,262 person-years of follow up (mean [SD] follow up, 19.5 [9.4], years). Details of the study population characteristics are provided in **Supplemental Table 4**.

The mean [SD] LE8, LE8 no sleep, LE8 health factor and LE8 health behavior sub- scores were 64.4 [14.2], 61.8 [15.8], 67.5 [19.6], and 61.3 [19.1], respectively, as shown in **Table 1**, stratified by index age group. All 4 health factor metrics had lower scores at higher ages, with the blood pressure metric showing the greatest decline (Young: 89.1, Middle Aged: 70.7, Older: 50.7) across age groups. Differences of health behavior scores across age groups varied for each metric; for example, nicotine exposure scores were highest (best) at the older ages (Young: 55.7 vs. Old: 65.0). Young and middle-aged participants had 20- and 10-point higher scores for health factors than behaviors (Young: 84.7 vs. 65.2; Middle Aged: 67.0 vs. 57.4), respectively, but older participants had higher behavior scores (health factor vs. health behavior: 61.5 vs. 64.8). The least favorable metrics among the young and middle-aged population were nicotine exposure and diet, and among the older participants they were blood pressure and diet. Sleep health was one of the most favorable heath behavior metrics in general and the scores were highest in the middle and older age groups.

**Table 1:** Descriptive Characteristics of Study Population. Values are expressed as mean (standard deviation) unless otherwise noted; results were calculated using 90 multiply imputed data sets.

|  | Young (20-39 years)<br>N= 4636 | Middle Aged (40-59 years)<br>N= 15982 | Older (60-79 years)<br>N= 12278 |
| --- | --- | --- | --- |
| Mean Age, yrs | 26.0 (3.7) | 51.7 (4.4) | 67.4 (5.5) |
| Male sex, % | 46.0% | 45.0% | 45.8% |
| Black race, % | 47.5% | 23.2% | 15.7% |
| < high school, % | 3.3% | 17.2% | 24.8% |
| High School, % | 16.4% | 28.7% | 27.7% |
| Some College, % | 32.4% | 17.2% | 20.4% |
| College and Higher, % | 48.0% | 36.8% | 27.2% |
| Mean LE8 health factor sub-scores (0-100) | 84.7 (14.8) | 67.0 (19.1) | 61.5 (17.9) |
| • Body mass index | 83.4 (26.2) | 66.2 (30.7) | 69.0 (28.5) |
| • Blood lipid | 80.1 (26.2) | 55.4 (31.6) | 54.2 (30.3) |
| • Blood pressure | 89.1 (20.7) | 70.7 (32.5) | 50.7 (35.3) |
| • Blood glucose | 86.1 (23.5) | 75.8 (26.4) | 72.3 (26.9) |
| Mean LE8 health behavior sub-scores (0-100) | 65.2 (18.4) | 57.4 (18.9) | 64.8 (18.7) |
| • Nicotine exposure | 55.7 (42.3) | 49.3 (40.6) | 65.0 (36.6) |
| • Physical activity | 83.4 (31.6) | 58.2 (45.0) | 65.1 (42.4) |
| • Sleep health | 79.2 (27.0) | 82.5 (27.2) | 83.0 (26.7) |
| • Diet | 42.4 (31.5) | 39.9 (31.3) | 46.1 (31.8) |
| Overall LE8 score -no sleep (0-100) | 74.3 (13.5) | 59.3 (15.5) | 60.3 (14.9) |
| Overall LE8 score (0-100) | 74.9 (12.3) | 62.2 (13.9) | 63.2 (13.5) |
| Overall LS7 score (0-14) | 10.8 (2.0) | 8.3 (2.3) | 8.4 (2.3) |

**Table 2A.**
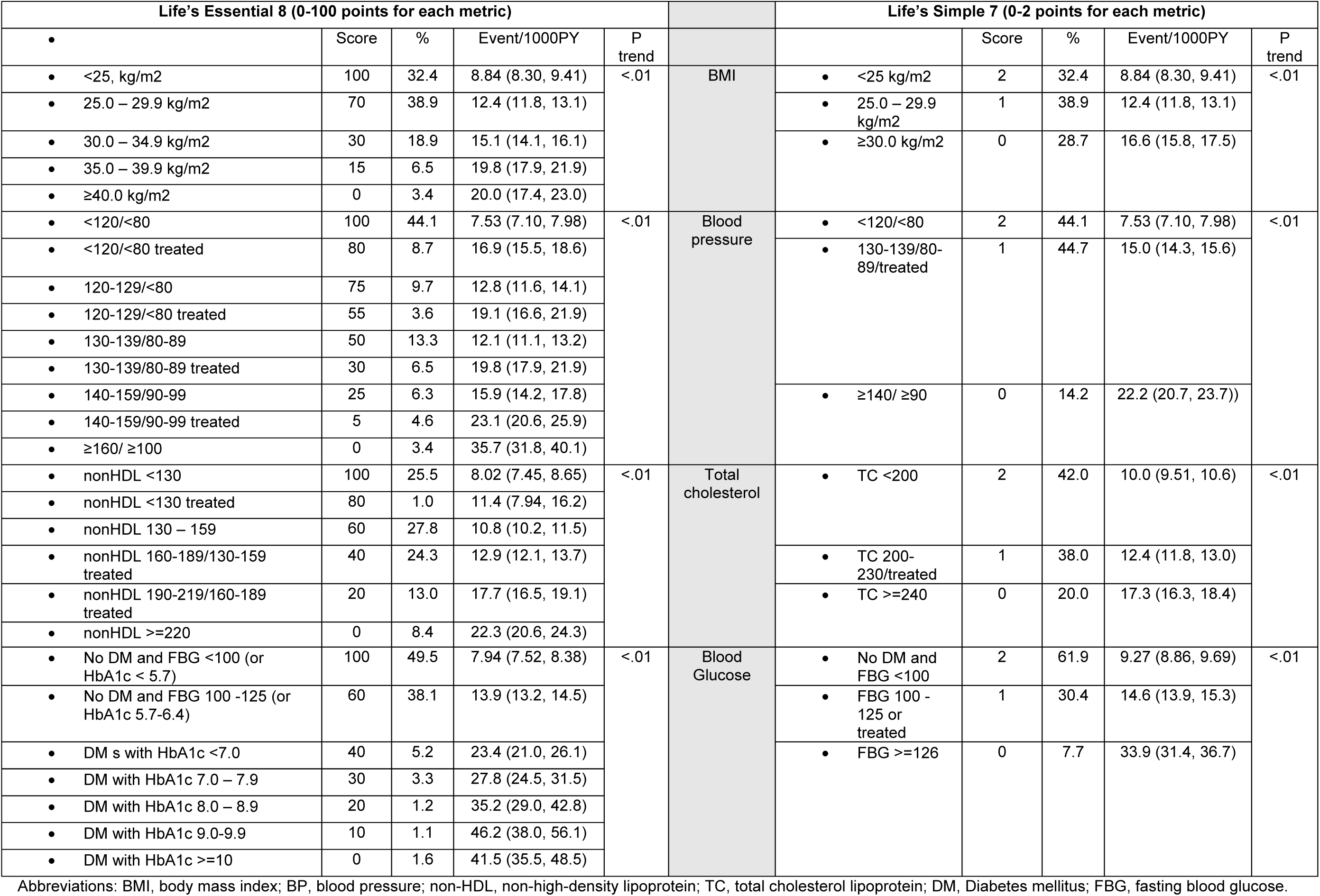
LE8 vs LS7 scores across health factors at Middle aged Group.

**Table 2B.**
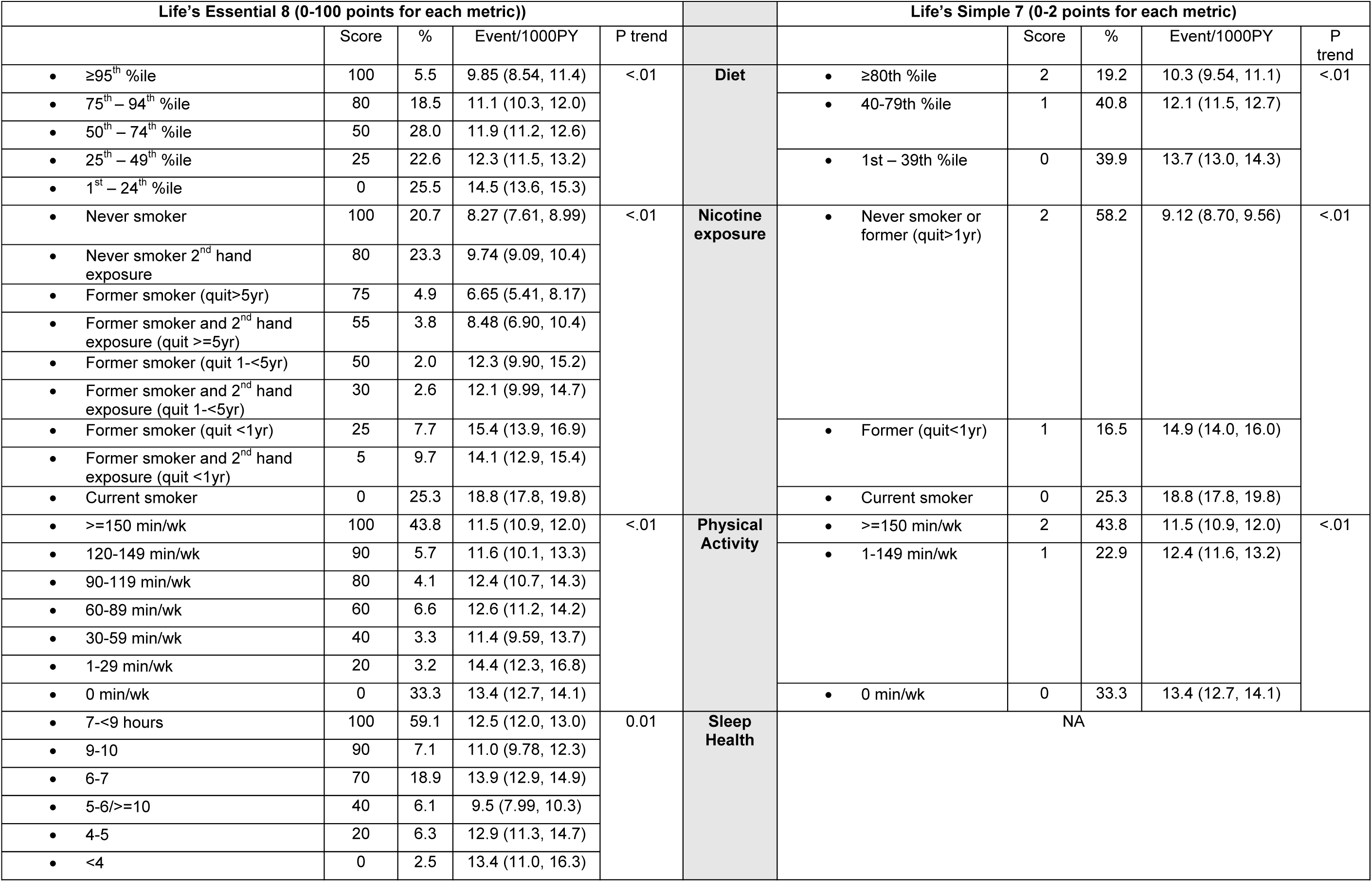
LE8 vs LS7 scores across health behaviors at Middle aged Group.

Inclusion of sleep as the eighth metric improved the LE8 score modestly at all ages (LE8 vs. LE8 no sleep - Young: 74.9 vs 74.3; Middle: 62.2 vs. 59.3; Old: 63.2 vs. 60.3). In Cox proportional models that evaluated the association of sleep health in relation to CVD risk, longer sleep was significantly associated with lower CVD risk and the association remained with the adjustment for LS7 or LE8 no sleep score respectively in middle-aged and older participants (results not presented).

The distribution of LE8 scores differed by sex, race, and education, with higher LE8 scores, including health factors and behaviors, in women, White, and higher educated participants across all age groups (**Supplemental Figure 1**). Education showed a strong dose response with LE8 score at all ages. We observed similar findings across White and Black subgroups.

Higher LE8 was significantly associated with CVD events across all ages after adjustment for age, self-reported gender and race, and education level (**Figure 1**). Each 10- points higher LE8 score was significantly associated with lower CVD risk and the strength of association was greatest among the young participants (HR (95%CI): Young 0.61 (0.56-0.66); Middle-aged 0.69 (0.67-0.71); Older: 0.77 (0.75-0.79)). The LE8 health factor score had a stronger association with CVD risk than the LE8 behavior score. We performed the same analyses based on different periods of follow-up (10-, 20- and 30-year) separately in sensitivity analyses, and the findings were consistent (results not presented). The overall LE8 score with sleep had slightly but not significantly stronger associations with CVD risk compared to the LE8 score without sleep (HR (95%CI): Young 0.62 (0.57-0.67); Middle 0.71 (0.67-0.72); Old: 0.78 (0.77-0.80)).

**Figure 1:**
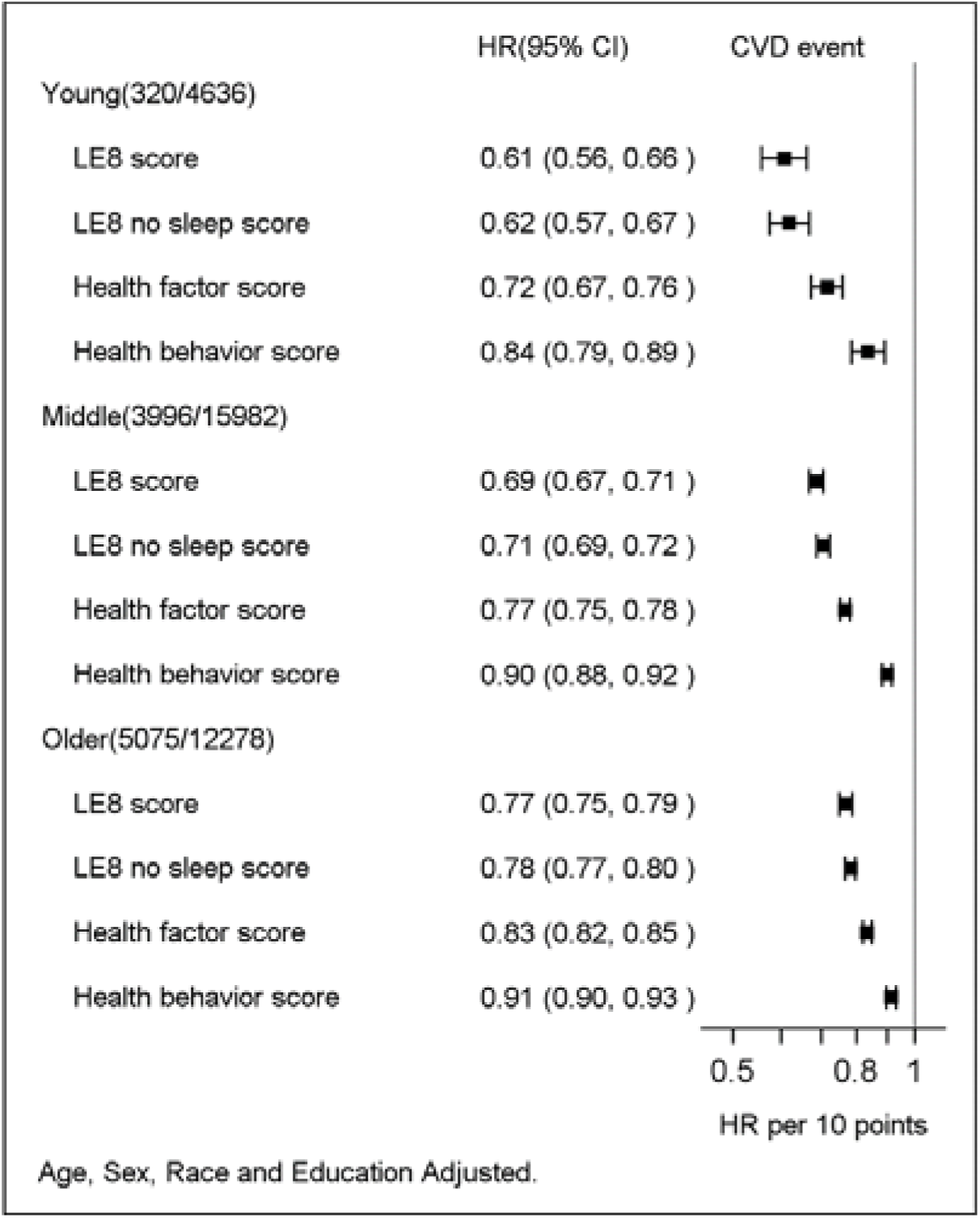
LE8 CVH score and hazards for cardiovascular events, for each 10 points higher LE8 score, stratified by age subgroups of the study population.

Associations between LE8 scores and subtypes of CVD events or all-cause mortality are displayed in **Supplemental Table 5**. Higher overall LE8, LE8 health factor and LE8 health behavior sub-scores were all significantly associated with lower risks for CHD, heart failure, stroke, CVD death and all -cause mortality across all age groups. Associations were generally strongest with CHD and weaker with all-cause mortality. Every 10-point higher LE8 score was significantly related with 43%, 35% and 25% lower CHD risk for young, middle, and older age groups respectively. LE8 health factor sub-scores had stronger associations with CHD and heart failure, but health behavior sub-scores had the stronger associations with all-cause mortality. Findings were consistent across all age groups.

For each individual CVH metric, the prevalence of all levels of LE8 and LS7 score category and their related CVD incidence rates were calculated and compared. For the sake of sufficient sample size and reliable risk estimation, we present the results for middle aged participants in Tables **2A and 2B.** For blood pressure, a level of ≥140/ ≥90 mm Hg was assigned a score of 0 in the LS7 algorithm, and these participants had a CVD incidence rate of 22.2 per 1000 person-year. This group, around 14.2% of the study population, was further stratified into three subgroups in the LE8 score system (score 25: 140-159/90-99 mm Hg (6.3%); score 5: 140-159/90-99 mm Hg treated (4.6%); and score 0: ≥160/ ≥100 mm Hg (3.4%)); the incidence of CVD increased in a dose dependent trend across these three groups (15.9 vs. 23.1 vs. 35.7, per 1000 person-years, respectively). Similarly, more than half of the participants were categorized as being never smokers or former smokers (quit>1yr) and universally assigned a score of 2 in LS7, but they were stratified into five subgroups with a wide range of incident CVD (6.7-12.3 per 1000 person-years) in the LE8 score algorithm. We observed similar findings across health factor and behavior metrics, with greater dose response and more precise levels of risk associated with the LE8 algorithm than the LS7 algorithm.

The concordant/discordant status across index age group is presented in **Table 3**. The agreement between the quartile rankings of LS7 and LE8 scores were 62.5% (kappa = 0.67) in young, 66.0% (kappa = 0.71) in middle aged, and 64.8% (kappa = 0.69) in older adults. More than three quarters of the top and bottom quartile remained unchanged as the concordant group, and half of the middle quartiles (25-50% and 50-75%) were reclassified upward or downward. The findings were similar across the age groups. The characteristics of reclassified participants are presented as **Supplemental Table 6**. The downward reclassified participants had universally lower overall LE8, LE8 no sleep, health factor and behavior sub-scores, respectively; and the obverse was true for the upward reclassified participants. The downward reclassified participants were more likely to report being younger, male, Black and less educated. We observed a greater health behavior sub-score difference across the reclassification group which indicates that these two CVH algorithms (LS7 vs. LE8) differ more on health behavior metrics. Taking the concordant group as the reference, the downward reclassified group had 6-10 points lower LE8 health behavior sub-scores (Young: 59.1 vs. 65.4; Middle: 47.2 vs. 57.1; Old: 54.7 vs. 64.7) and 2-3 points lower LE8 health factor sub-scores (Young: 82.2 vs. 84.4; Middle: 63.7 vs. 66.6; Old: 58.0 vs. 61.4). Lower sleep scores were observed in the downward reclassified group.

**Table 3:**
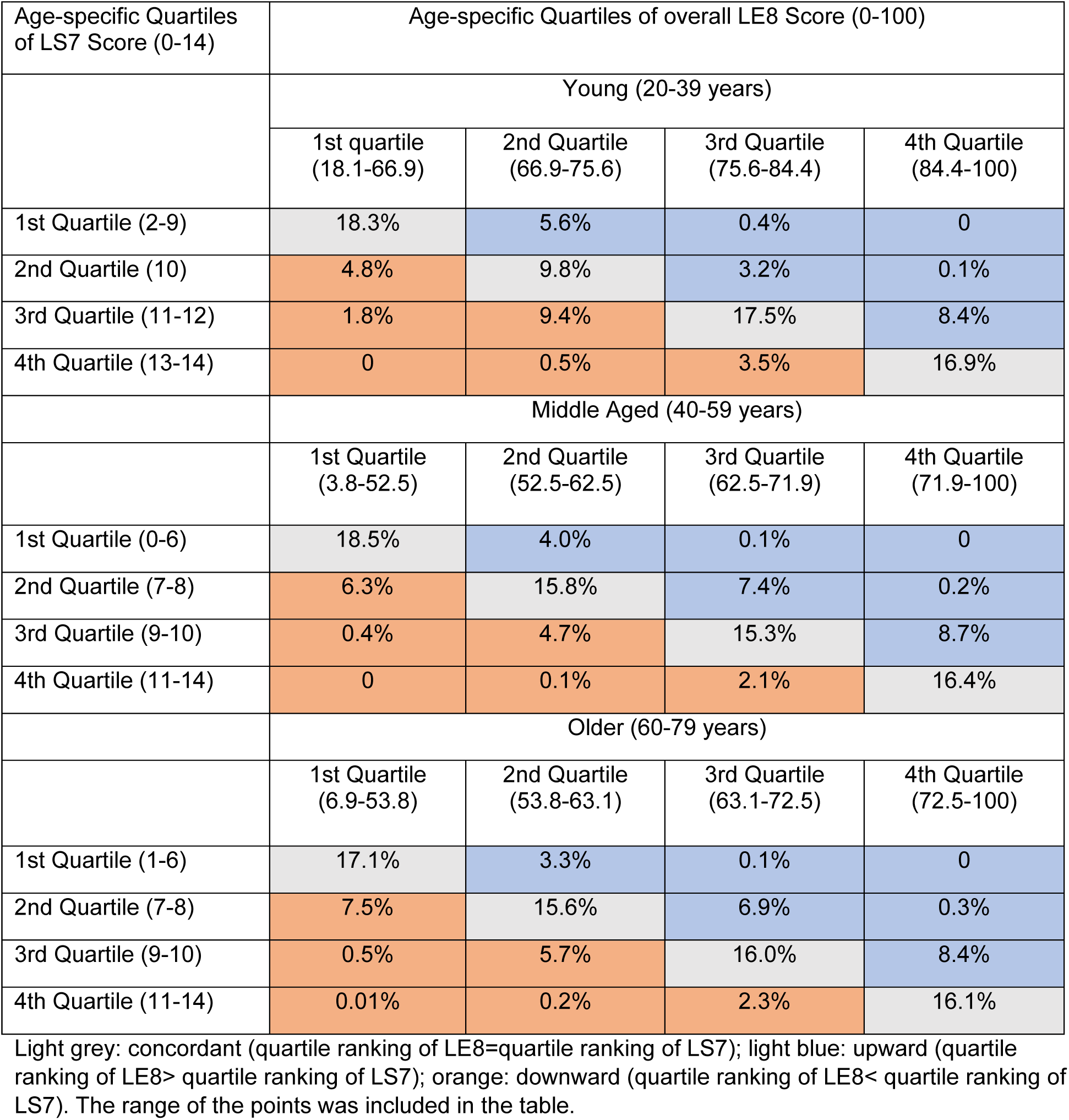
% of participants with LS7-LE8 reclassification across index age group.

| Age-specific Quartiles of LS7 Score (0-14) | Age-specific Quartiles of overall LE8 Score (0-100) |  |  |  |
| --- | --- | --- | --- | --- |
|  | Young (20-39 years) |  |  |  |
|  | 1st quartile<br>(18.1-66.9) | 2nd Quartile<br>(66.9-75.6) | 3rd Quartile<br>(75.6-84.4) | 4th Quartile<br>(84.4-100) |
| 1st Quartile (2-9) | 18.3% | 5.6% | 0.4% | 0 |
| 2nd Quartile (10) | 4.8% | 9.8% | 3.2% | 0.1% |
| 3rd Quartile (11-12) | 1.8% | 9.4% | 17.5% | 8.4% |
| 4th Quartile (13-14) | 0 | 0.5% | 3.5% | 16.9% |
|  | Middle Aged (40-59 years) |  |  |  |
|  | 1st Quartile<br>(3.8-52.5) | 2nd Quartile<br>(52.5-62.5) | 3rd Quartile<br>(62.5-71.9) | 4th Quartile<br>(71.9-100) |
| 1st Quartile (0-6) | 18.5% | 4.0% | 0.1% | 0 |
| 2nd Quartile (7-8) | 6.3% | 15.8% | 7.4% | 0.2% |
| 3rd Quartile (9-10) | 0.4% | 4.7% | 15.3% | 8.7% |
| 4th Quartile (11-14) | 0 | 0.1% | 2.1% | 16.4% |
|  | Older (60-79 years) |  |  |  |
|  | 1st Quartile<br>(6.9-53.8) | 2nd Quartile<br>(53.8-63.1) | 3rd Quartile<br>(63.1-72.5) | 4th Quartile<br>(72.5-100) |
| 1st Quartile (1-6) | 17.1% | 3.3% | 0.1% | 0 |
| 2nd Quartile (7-8) | 7.5% | 15.6% | 6.9% | 0.3% |
| 3rd Quartile (9-10) | 0.5% | 5.7% | 16.0% | 8.4% |
| 4th Quartile (11-14) | 0.01% | 0.2% | 2.3% | 16.1% |

Light grey: concordant (quartile ranking of LE8=quartile ranking of LS7); light blue: upward (quartile ranking of LE8> quartile ranking of LS7); orange: downward (quartile ranking of LE8< quartile ranking of LS7). The range of the points was included in the table.

Even after adjustment for the baseline LS7 score, reclassification by LE8 was independently associated with CVD risk and this association was stronger for older age participants. 16.3% of older participants were downward reclassified from a higher to a lower quartile from LS7 to LE8, and they had 31% higher CVD risk. On the other hand, 19% of older participants were upward reclassified (to a higher CVH score quartile), which was associated with 17% lower CVD risk (**Table 4**). Reclassification status had no significant association with CVD risk among the young group.

**Table 4.** Multivariable-Adjusted Cox proportional hazards models for Associations of LS7 and reclassification with Cardiovascular event across index age groups.

|  | Crude Incidence Rate per 1, 000 Person-Years | Adjusted Hazard Ratio (95% CI) |  |  |  |
| --- | --- | --- | --- | --- | --- |
|  |  | LS7 score (Per 1 point) | Concordant Group (Quartile ranking of LE8 = quartile ranking of LS7) | Upward reclassification (Quartile ranking of LE8 > quartile ranking of LS7) | Downward reclassification (Quartile ranking of LE8 < quartile ranking of LS7) |
| Young (20-39 years) | 2.17 (1.95, 2.43) | 0.72 (0.68, 0.76) | Ref | 0.92 (0.63, 1.34) | 0.96 (0.62, 1.49) |
| Middle Aged (40-59 years) | 12.3 (12.0, 12.7) | 0.80 (0.78, 0.81) | Ref | 0.87 (0.79, 0.96) | 1.20 (1.07, 1.35) |
| Older (60-79 years) | 29.6 (28.8, 30.4) | 0.86 (0.85, 0.87) | Ref | 0.82 (0.75, 0.91) | 1.31 (1.01, 1.24) |

Both LS7 scores and reclassification status were included in the model, and was further adjusted for age, sex, race/ethnicity, and education status; we imputed 90 datasets, and each of dataset was analyzed separately, and the results were pooled as the final imputed estimates using Rubin's rules.

## Discussion

In this analysis of more than 32,000 US adults (with 642,000 person-years of follow up), higher LE8 scores were associated with significantly lower risk for CVD events, CVD subtypes, and all-cause mortality. Each 10-point higher LE8 score was associated with 23-40% lower CVD risk across index ages. LE8 health factors, reflecting more proximal causal CVD risk factors had stronger associations with CVD and subtype CVD risk than health behaviors in general. Young and middle-aged participants had poorer health behavior scores compared to health factor scores. The updated LE8 point scoring system provided more granular estimates of related CVD risk compared to the LS7 score across each health and behavior metric. Reclassification from LS7 to LE8 was related to the health behaviors as well as health factors, and reclassification status (upward or downward) added significantly to the LS7 score in the association with CVD risk. This study underscores the implications of applying the more granular LE8 score for cardiovascular risk estimates, the importance of having high LE8 regardless of age, and the significance of improving behavior metrics among young and middle-aged individuals.

This study is the first to report on the association of AHA’s updated LE8 CVH score with the risks of CVD, subtype CVD and death in US adults. Some recent studies examined LE8 score with mortality data and suggested that improvement in health behaviors and factors may reduce the risk of dying from heart disease and total mortality ^10, 24^. Our study leveraged a large, harmonized dataset and examined fatal and nonfatal CVD events across the life course with a long follow-up period. We observed that LE8 scores, both with and without sleep, had significant associations with CVD, and there were stronger associations between LE8 health factor sub-scores and CVD risk compared to LE8 health behavior sub-scores across all age groups. Bias towards health behaviors measures of engagement may lead to greater measurement error which can attenuate the associations. The findings also applied over medium- and long-term follow-up periods. Given that health behaviors are on the causal pathway leading to health factors and ultimately CVD events, improving health behaviors will result in overall improvements in CVH health. Of note, younger participants had greater behavior score differences across sociodemographic subgroups and the health behavior score was 20- points lower than the health factor score overall. This observation underscores the significance of promoting healthy behaviors to improve population CVH status, especially among younger and socioeconomically disadvantaged groups. Abstinence from tobacco, healthier diet, or 30 minutes more exercise every week would help the young participants achieve 20 or more metric points of health behavior score which could ultimately be associated with ∼30% lower CVD risk.

The benefits of optimal LS7 for healthier aging and CVD risk reduction have been investigated across diverse population groups ^12, 13, 25, 26^. LE8 was developed as a tool to improve assessment of CVH using updated and modified LS7 metrics with inclusion of sleep as the eighth metric. Our study demonstrates that the new LE8 algorithm has more sensitivity to capture inter-individual differences with direct implications for CVD risk. Compared to the LS7 score, each component of LE8 score has more levels and that allows each level to have a narrower range of values and increases the sensitivity of the score. The LS7 score doesn’t quantify the metrics with the respective measurement beyond the traditional clinical categories whereas the LE8 score evaluates CVH using the entire range of the respective CVH metric. Therefore, BMI, blood lipids, blood glucose, blood pressure in LE8 score reflect the incremental increase in CVD risk with these risk factors beyond the clinical range value. The LE8 scoring system accounts for medication usage and assigns 20 points deduction for those using medication to achieve a given level, which appears to be a more appropriate approach, reflecting greater cumulative exposure to hypertension or hyperlipidemia. Our results showed a higher incidence of CVD among those medication users compared to untreated peers with similar CVH levels. Furthermore, the association of the secondhand smoke with the increased risk of CVD event is accounted in the LE8 score ^27^.

Furthermore, sleep health was an important factor affecting health ^4, 28–30^. Our study found that sleep health was associated with CVD risk and this significant relationship remained with LS7 and LE8 no sleep score adjustment respectively, which implied that sleep health has an independent relationship with CVD risk in addition to the other 7 metrics. Of interest, we did not observe a dose-response trend of CVD incidence rates across the sleep points that was observed in other LE8 metrics. The participants with 5-6 or >=10 sleep hours had the lowest CVD rates compared to the others. We found that this group included more higher-educated white females and they achieved more optimal CVH metrics except long sleeping (>=10). The mechanism of this finding is not entirely clear, and more studies are needed to replicate these findings and investigate potential reasons for them.

Previous studies have shown the dose response linear relationship of LE8 score and CVD risk^10, 24^. In this study, we ranked the participants by LE8 and LS7 score quartiles and examined reclassification by quartiles from LS7 to LE8. We found that downward reclassification from LS7 to LE8 score was associated with higher CVD risk; for example, downward reclassified participants had a 31% higher CVD risk among older participants. Adults aged 65 and older account for 82% of all death attributable to heart disease, and CVD in older Americans imposes a huge burden in terms of mortality and medical costs ^31^. The implementation of LE8 scoring among older participants may help to distinguish the highest-risk older participants and assist primary prevention decisions.

Our study has several important strengths including the being the first study to report the association between LE8 and CVD risk in a US cohort population across the life course, and the long follow up of the cohort (∼20 years) thus allowing us to examine the association with events even for younger individuals. However, several limitations deserve mention. First, although we adjusted for potential confounders, residual confounding may be present. Because of limited availability, we were unable to assess other social determinants (e.g., family income and relative poverty index) and psychosocial factors (e.g., depression). Second, we used baseline CVH assessments, and it may be interesting to investigate further how the longitudinal change of LE8 score influences CVD events and health outcomes. Third, the analysis is limited to Black and White adults. The association of LE8 with CVD events experienced by Hispanic, Asian and Native American population are not evaluated. However, prior studies of LS7 have suggested similar benefits of higher CVH for all race/ethnic groups. Fourth, self-reported total sleep time tends to be less precise than objectively measured sleep time ^32^, which may bias our findings. Fifth, sleep health and HBA1c were heavily imputed using the observed records. The imputed data reflect the association with the observed characteristics, but the unobserved characteristics (shifting work schedule, smartphone addiction) ^33, 34^ could alter or influence the association.

In conclusion, Life’s Essential 8 was significantly associated with risks for CVD, subtype CVD events and all-cause mortality among young, middle-aged, and older- aged participants across sociodemographic groups. Compared with the LS7 algorithm, LE8 had a more refined, dose-response association with CVD risk and higher sensitivity for inter-individual variance. Communication and health promotion strategies for improving healthy lifestyle should be expanded from nicotine exposure, diet, and physical activity to include sleep health. Other health-promoting activities to improve healthy diet, exercising and early abstinence of tobacco should assist in achieving optimal CVH status, with attendant lower individual and population burden of CVD. Likewise, vigorous clinical measures to control adverse levels of CVH factors (hypertension, dyslipidemia, dysglycemia, overweight and obesity) must continue. Measures that advocate for optimal LE8 should be encouraged beginning from young ages and extending throughout the life course.

## Data Availability

the data supporting the findings of this study are available via Biolincc.

https://biolincc.nhlbi.nih.gov/home/

